# Obesity Intervention in Sub-Saharan Africa: A systematic review protocol

**DOI:** 10.1101/2023.07.18.23292826

**Authors:** Kingsley Agyemang, Nana Anokye

## Abstract

**Introduction:** The increasing prevalence of obesity in Sub-Saharan Africa (SSA) has made it a pressing public health issue that requires effective interventions tailored to the region’s unique challenges. This protocol outlines a systematic review approach that aims to examine the existing evidence on obesity interventions in SSA, focusing on their effectiveness, implementation strategies, and potential barriers.

**Methods and analysis:** To identify relevant studies in Sub-Saharan Africa (SSA), MEDLINE (PubMed), MEDLINE (EBSCOHost), Scopus, Web of Science, PsycINFO, Cochrane and EMBASE databases will be searched during the review. Studies on obesity interventions in SSA will be considered for inclusion criteria, encompassing various intervention types, target populations, and study designs. Abstracts and titles will be independently assessed by two reviewers, who are aware of the author and journal information. A third reviewer will be consulted for cases where agreement was not reached or when clarity will be needed. The methodological quality of the included research will be assessed using the “Effective Public Health Practise Project quality assessment tool for quantitative studies. For data synthesis and reporting, two independent reviewers will assign effect scores to each intervention’s outcome category (dietary behaviour, physical activity, or anthropometric outcomes) through a duplicate scoring exercise.

**Ethics and dissemination:** Since no original data will be collected as part of this review, ethical approval is not required. The completed review will be submitted for publication in a peer-reviewed journal and presented at conferences.

**PROSPERO registration number:** CRD42023430503

## Introduction

### Rationale

Obesity is an ongoing pandemic[1]–[3] and following changes in dietary patterns and physical activity, the prevalence of obesity is anticipated to increase in both developed and developing nations[1], [4]–[7]. This poses a significant healthcare issue, due to its link to chronic diseases like diabetes, cardiovascular diseases, and certain types of cancer [8]. Over the years, Obesity has been described as “the silent killer” [9], further emphasizing its grave implications for public health. Obesity is defined by an elevated body mass index (BMI) and/or an excess accumulation of body fat[10]–[12]. Between 1975 and 2016, the worldwide occurrence of obesity experienced an almost threefold increase[3], [13], [14], with most of the world’s population living in nations where being overweight or obese kills more people than being underweight [15].

The World Health Organization reported that in 2016, there was a prevalence of over 650 million obese adults, 340 million individuals aged 5 to 19 with obesity, and approximately 39 million children under the age of 5 affected by obesity [15]. However, the World Obesity Federation has issued a warning, predicting that if immediate action is not taken, the number of obese adults is projected to reach 1 billion by 2030[14], [16]. Additionally, economies of most low-middle-income countries including Sub-Sahara African countries are further affected as obesity has a heavy toll on productivity, increasing disease in sole wage workers[17]–[21].

It is estimated that Overweight and Obesity (OAO) cost 2.19% of the global Gross Domestic Product (GDP), with an average per capita value of US$20 in Africa. This is expected to increase 12-25 times in LMICs including Africa if immediate intervention is introduced to address this issue [17]. Literature has shown that the rates of overweight and obesity are increasing in sub-Saharan Africa (SSA), particularly among urban populations[22]–[27]. This is further established in a multi-country cross-sectional study conducted in some sub-Saharan African countries recorded overweight prevalence of 31% and obesity prevalence of 34%[27]. It is important to note that the prevalence and risk factors of obesity differ across various regions globally[28], in sub-Saharan Africa, various factors such as socioeconomic status, age, parity, marital status, physical inactivity, body weight perceptions, and increased energy levels have been identified as significant predictors of overweight and obesity [29]. To effectively address this challenge in the SSA region, it is crucial to develop and adopt obesity interventions that are tailored to the specific characteristics of the population, recognizing the variations in demographics, socio-economic factors, and contextual factors [30]–[32].

Several interventions are currently used to manage weight in cases of obesity, which usually focused on implementing lifestyle changes that include physical activity and dietary recommendations[9], [33]–[35]. Nevertheless, there are mixed evidence on the effectiveness of these interventions[7], [36]–[40] which remains a major challenge in the fight against obesity especially in SSA. Additionally, there is paucity of studies on the obesity and the effectiveness of the available obesity interventions in SSA[38], [41]–[46]. Therefore, evidence from other countries and settings cannot be used to assume the best avenues for interventions in SSA [47] hence the need for robust evidence from systematic reviews assessing the effectiveness of the interventions in the context in which it was implemented, which may be culturally appropriate, cost-effective, and long-lasting public health preventive measure[15], [48]–[50].

Our scoping review reveals that no systematic review has been conducted in SSA to provide evidence on the effectiveness of obesity interventions introduced to reduce obesity in the region. However, the available systematic review on obesity intervention in SSA, focussed on childhood obesity[32] but the rise in obesity rates is concerning globally, affecting all age groups and socioeconomic categories. The World Health Organization’s (WHO) Global Action Plan encourages national governments to create public health strategies and NCD including obesity targets to enhance the health of the population [51], hence this systematic review will provide evidence on the effectiveness of several obesity interventions in SSA to inform policy direction. This systematic review will therefore be the first to address this gap in knowledge through the evaluation of the effectiveness of interventions targeting obesity in Africa by assessing existing evidence and considering diverse contexts. The review seeks to inform public health practice and guide the development of evidence-based interventions to combat obesity in Africa.

## Objectives

The review will:

1. To conduct a thorough and systematic review of the existing literature on obesity interventions in Sub-Saharan Africa.
2. To evaluate the effectiveness of interventions aimed at addressing obesity in Sub-Saharan Africa, encompassing both lifestyle interventions and other approaches.
3. To find areas for further research and identify gaps in the current evidence base regarding obesity interventions in Sub-Saharan Africa.
4. To provide evidence-based recommendations for interventions that can effectively tackle the obesity epidemic in Sub-Saharan Africa while taking into account the region’s unique cultural, socio-economic, and healthcare system factors.

## METHODS

The reporting of this systematic review protocol follows the guidelines outlined in the Preferred Reporting Items for Systematic Reviews and Meta-Analyses Protocol (PRISMA-P) statement[52]. The PRISMA checklist can be found in Supplementary file 1. Since this review focuses on published studies, obtaining ethics approval is not necessary. The review has been registered with the International Prospective Register of Systematic Reviews (PROSPERO) under the registration number CRD42023430503, with the registration date being 19th June 2023.

## Eligibility criteria Inclusion criteria

### Study setting

Sub-Saharan African nation in accordance with the regional criteria by the world bank[53]

### Population

1. Studies were included if interventions targeted participants who were obese or overweight, regardless of age, including both children and adults. 2. Interventions carried out in community-based settings, which may include but are not limited to schools, workplaces, and recreational centres.

### Intervention

Any intervention to reduce or prevent obesity/overweight in any Sub-Sahara African country, including but not limited to dietary, physical activity, lifestyle weight management and behavioural change techniques.

### Comparator

1. Studies that may have compared different interventions targeting obesity in community-based settings. 2. In certain cases, studies exploring the effectiveness of specific interventions may have included placebo or sham interventions as controls.

### Outcomes

1. Body composition, as well as outcomes associated to adiposity, such as the prevalence of overweight and obesity. 2. Intermediate behavioural outcomes, including modifications in sedentary behaviour, eating behaviour, and physical activity and fitness levels. 3. In terms of behavioural outcomes, both objective and subjective assessments of physical activity, dietary behaviour, and other relevant behaviours such as sedentary behaviour are considered appropriate.

### Study design

Experimental studies, which consisted of randomized controlled trials (RCTs), quasi-experimental studies, controlled clinical trials, and cluster trials. prospective cohort studies with a control group, interrupted time series and repeated measure studies, and natural experiments.

### Exclusion criteria

1. Interventions specifically targeted participants who were not overweight or obese, regardless of age, including both children and adults.
2. Studies aimed at expanding interventions in medical settings like hospitals or general practice.
3. Any intervention that involved using medication, surgery, or dietary supplements to help people lose weight.
4. Interventions that focused on populations with pre-existing medical conditions or obesity-related co-morbidities.
5. Non-experimental studies, cross-sectional studies, non-human studies, laboratory-based studies.
6. Health outcomes that do not report on relevant adiposity outcomes.
7. Behavioural treatments that do not include important behavioural outcomes such as increased in physical activities, dietary behaviour, fitness, or sedentary behaviour.
8. Working papers, reports, dissertations, books, websites, conference abstracts, and research protocols.
9. Studies that were conducted in countries outside the sub-Saharan region.

### Search strategy

A scoping review of systematic review methodologies in the discipline was used to choose databases for this study[32], [38], [54]–[57]. To identify potentially eligible studies, an extensive search will be conducted across multiple databases including MEDLINE (PubMed), MEDLINE (EBSCOHost), Scopus, Web of Science, PsycINFO, Cochrane and EMBASE. Subject header and free text searches will be performed, employing Boolean search methods like “AND” and “OR” aligning with the eligibility criteria for the systematic review. Unique search terms will be employed for each database, tailoring the search strategy accordingly. The comprehensive search strategy of PubMed, including the specific details, can be found in the Supplementary material (2). Our search will not be limited by language or publication year.

### Study selection

Abstracts and titles will be independently assessed by two reviewers, who were aware of the author and journal information. Google Translate will be utilized to determine the eligibility of abstracts not published in English. Full papers of potentially eligible studies will be obtained for a more thorough evaluation. For manuscripts not published in English, translation of the entire text will be done. The two reviewers reached a consensus on study inclusion, and a third reviewer will be consulted for cases where agreement was not reached or when clarity was needed. Manuscripts with incomplete texts will be excluded based on the primary reason stated.

### Data management and extraction

A pretested spreadsheet will be used by one reviewer to extract the data, while a second reviewer cross-checked the generated data and added any missing information. The data that will be extracted included a variety of information, such as the study title, intervention details, target population, study design, information on comparisons or control groups, outcomes, measurement methods, publication type, publication year, study setting, country, language, inclusion criteria, baseline descriptive data, randomization procedure, intervention duration, follow-up period, number of follow-ups, participant attrition, sample size, and effectiveness for relevant outcome statistical tests and adjustments, subgroup effects (if applicable), and any additional publications referenced in the article[32].

### Risk of bias and quality assessment

The methodological quality of the included research will be assessed using the “Effective Public Health Practise Project quality assessment tool for quantitative studies”[58]. This tool evaluated six criteria, including selection bias, research design, confounders, blinding, data collection techniques, and withdrawals/drop-outs. Each criterion will be rated as weak, moderate, or strong for each study. The overall score for each study will be calculated by summing the ratings across the six components. Studies that received no weak ratings were categorized as strong, those with one weak rating will be categorized as moderate, and those with two or more weak ratings will be categorized as weak[55].

### Data synthesis and reporting

Two independent reviewers will assign effect scores to each intervention’s outcome category (dietary behaviour, physical activity, or anthropometric outcomes) through a duplicate scoring exercise. This scoring method, employed in a previous review[32], [59], [60], involved assigning scores ranging from “++” to “--” based on the composite of relevant outcomes reported in each study. A score of “++” will indicate a statistically significant and clearly intervention-attributable desired change on the primary outcome or most outcomes of interest. A score of “+” will indicate a desired change on the primary outcome or mostly desired changes on relevant outcomes. A score of “0” will indicate no changes, mostly no changes, or both positive and negative changes. A score of “-” will signify a statistically significant, obviously intervention-attributable negative change on the primary outcome or most outcomes of interest, while “--” denotes a statistically significant, definitely intervention-attributable negative change on the primary outcome or most outcomes of interest. The two authors will review scoring discrepancies, ensuring that each intervention aligns with the established scoring criteria. Additionally, they will create summary scores for each behaviour by comparing the number of varied scores and assigning the most frequent score as the summary score.

### Study status

In April 2023, a scoping study of the obesity interventions in Africa was conducted to inform the techniques of the present review The scoping review involved identifying existing systematic reviews on obesity interventions and identifying knowledge gaps. However, the results of the scoping review have not been submitted for publication. In May 2023, the review design in SSA was proposed, and methods were established. In June 2023, the search strategy was tried, and the protocol was also created. In July 2023, the protocol was submitted for peer review for the first time. Data collection will begin after the protocol for publication is approved and will be finished in three months.

### Patient and public involvement

The involvement of patients and the public is not included in this study since original data collection will not take place. The study will adhere to established best practice guidelines for conducting systematic reviews[61], [62].

## DISCUSSION

The systematic review protocol aims to improve our understanding of obesity interventions in Sub-Saharan Africa. By reviewing the literature systematically, assessing the effectiveness of interventions, identifying gaps in knowledge, and providing evidence-based recommendations, this study will contribute to the existing knowledge and offer valuable insights for addressing the obesity epidemic in this region. The results of this review will be shared widely to ensure their reach and impact. The insights obtained from this study will be disseminated through conference presentations and published in peer-reviewed journals, making them accessible to the broader research community and stakeholders.

### Strength and limitations of the study

- This is the first systematic review on obesity interventions in SSA that aims to provide quality evidence for the effectiveness of obesity interventions in Sub-Sahara Africa and identify salient features of effective interventions.
- The study aims to suggest evidence-based interventions that suit the region by considering the specific factors of culture, economy, and health system in Sub-Saharan Africa.
- The study aims to identify gaps in the current evidence base, highlighting areas that require further research and investigation.
- We did not exclude any articles based on language, but we searched only in English. This might have limited our search results if some articles in other languages did not have abstracts or keywords in English.
- Publication bias might affect this review because it only includes studies that have been peer reviewed.

## Data Availability

In this systematic review protocol, we carefully evaluated data availability. The data to be utilized in this study were obtained from credible databases or, when necessary, from the authors of the included research. We affirm that all data that will be described in the publication will be available for examination upon request, and we are dedicated to maintaining transparency and reproducibility in our research.

## Acknowledgements

We express our gratitude to the Brunel Global Health Academy for their valuable support and assistance.

## Author Contributions

The concept for the review was developed by NA and KA. KA took the lead in writing the initial draft, while both KA and NA collaborated in revising the protocol. NA will assume the role of being the guarantor of the review, ensuring its overall integrity and accuracy.

## Patient consent for publication

Not required.

