## Supplementary material 2 for "Obesity Intervention in Sub-Saharan Africa: A systematic review protocol"

| Search Term 1 | "Obesity"[MeSH Terms] OR "body mass index"[MeSH Terms] OR "Body Weight"[MeSH Terms] OR ("overweight"[MeSH Terms] OR "overweight"[All Fields] OR "overweighted"[All Fields] OR "overweightness"[All Fields] OR "overweights"[All Fields]) OR "obes*"[All Fields] OR ("obeses"[All Fields] OR "Obesity"[MeSH Terms] OR "Obesity"[All Fields] OR "obese"[All Fields] OR "obesities"[All Fields] OR "obesity s"[All Fields]) OR "bmi"[All Fields] OR "body mass"[All Fields] OR (("human body"[MeSH Terms] OR ("human"[All Fields] AND "body"[All Fields]) OR "human body"[All Fields] OR "body"[All Fields]) AND ("molecular weight"[MeSH Terms] OR ("molecular"[All Fields] AND "weight"[All Fields]) OR "molecular weight"[All Fields] OR "mass"[All Fields])) OR "body mass index"[All Fields] OR "body mass index"[All Fields] OR "Body Weight"[All Fields] OR ("Body Weight"[MeSH Terms] OR ("body"[All Fields] AND "weight"[All Fields]) OR "Body Weight"[All Fields]) OR "body composition"[All Fields] OR ("weight s"[All Fields] OR "weighted"[All Fields] OR "weighting"[All Fields] OR "weightings"[All Fields] OR "weights and measures"[MeSH Terms] OR ("weights"[All Fields] AND "measures"[All Fields]) OR "weights and measures"[All Fields] OR "weight"[All Fields] OR "Body Weight"[MeSH Terms] OR ("body"[All Fields] AND "weight"[All Fields]) OR "Body Weight"[All Fields] OR "weights"[All Fields]) OR "weight status"[All Fields] OR "body size"[All Fields] OR "fatness"[All Fields] OR "body fat"[All Fields] OR "adipos*"[All Fields] OR "nutritional status"[All Fields] OR ("nutritional status"[MeSH Terms] OR ("nutritional"[All Fields] AND "status"[All Fields]) OR "nutritional status"[All Fields]) |
| --- | --- |
| Search Term 2 | "obesity intervention"[All Fields] OR "obesity treatment"[All Fields] OR "weight management"[All Fields] OR "obesity management"[All Fields] OR "obesity control"[All Fields] OR "health promotion"[All Fields] OR "health education"[All Fields] OR "weight loss"[All Fields] OR "lifestyle modification"[All Fields] OR "dietary intervention"[All Fields] OR "physical activity intervention"[All Fields] OR "recreation*"[All Fields] OR "physical activ*"[All Fields] OR ("exercise"[MeSH Terms] OR "exercise"[All Fields] OR ("physical"[All Fields] AND "activity"[All Fields]) OR "physical activity"[All Fields]) OR "sport*"[All Fields] OR ("sport s"[All Fields] OR "sports"[MeSH Terms] OR "sports"[All Fields] OR "sport"[All Fields] OR "sporting"[All Fields]) OR ("youth sports"[MeSH Terms] OR ("youth"[All Fields] AND "sports"[All Fields]) OR "youth sports"[All Fields] OR ("youth"[All Fields] AND "sport"[All Fields]) OR "youth sport"[All Fields]) OR "physical education"[All Fields] OR ("physical education and training"[MeSH Terms] OR ("physical"[All Fields] AND "education"[All Fields] AND "training"[All Fields]) OR "physical education and training"[All Fields] OR ("physical"[All Fields] AND "education"[All Fields]) OR "physical education"[All Fields]) OR "physical training"[All Fields] OR "exercis*"[All Fields] OR ("exercise"[MeSH Terms] OR "exercise"[All Fields] OR "exercises"[All Fields] OR "exercise therapy"[MeSH Terms] OR ("exercise"[All Fields] AND "therapy"[All Fields]) OR "exercise therapy"[All Fields] OR "exercising"[All Fields] OR "exercise s"[All Fields] OR "exercised"[All Fields] OR "exerciser"[All Fields] OR "exercisers"[All Fields]) OR "energy expenditure"[All Fields] OR ("energy metabolism"[MeSH Terms] OR ("energy"[All Fields] AND "metabolism"[All Fields]) OR "energy metabolism"[All Fields] OR ("energy"[All Fields] AND "expenditure"[All Fields]) OR "energy expenditure"[All Fields]) OR "physical inactivity"[All Fields] OR ("sedentary behavior"[MeSH Terms] OR ("sedentary"[All Fields] AND "behavior"[All Fields]) OR "sedentary behavior"[All Fields] OR ("physical"[All Fields] AND "inactivity"[All Fields]) OR "physical inactivity"[All Fields]) OR "physical fitness"[All Fields] OR "active travel"[All Fields] OR ("sedentaries"[All Fields] OR "sedentariness"[All Fields] OR "sedentary"[All Fields]) OR ("motor activity"[MeSH Terms] OR ("motor"[All Fields] AND "activity"[All Fields]) OR "motor activity"[All Fields]) OR ("physical exertion"[MeSH Terms] OR ("physical"[All Fields] AND "exertion"[All Fields]) OR "physical exertion"[All Fields]) OR "physical education and training"[All Fields] OR "physical activity environment"[All Fields] OR ("fitness"[All Fields] OR "fitnesses"[All Fields]) OR ("fitness"[All Fields] OR "fitnesses"[All Fields]) OR ("inactivities"[All Fields] OR "sedentary behavior"[MeSH Terms] OR ("sedentary"[All Fields] AND "behavior"[All Fields]) OR "sedentary behavior"[All Fields] OR "inactivity"[All Fields]) OR "diet intervention"[All Fields] OR "dietary intake"[All Fields] OR ("eating"[MeSH Terms] OR "eating"[All Fields] OR ("dietary"[All Fields] AND "intake"[All Fields]) OR "dietary intake"[All Fields]) OR "dietary behavior*"[All Fields] OR "dietary behaviour*"[All Fields] OR ("eating"[MeSH Terms] OR "eating"[All Fields]) OR ("eating"[MeSH Terms] OR "eating"[All Fields]) OR "diet*"[All Fields] OR ("diet"[MeSH Terms] OR "diet"[All Fields]) OR "nutrition*"[All Fields] OR ("nutrition s"[All Fields] OR "nutritional status"[MeSH Terms] OR ("nutritional"[All Fields] AND "status"[All Fields]) OR "nutritional status"[All Fields] OR "nutrition"[All Fields] OR "nutritional sciences"[MeSH Terms] OR ("nutritional"[All Fields] AND "sciences"[All Fields]) OR "nutritional sciences"[All Fields] OR "nutritional"[All Fields] OR "nutritionals"[All Fields] OR "nutritions"[All Fields] OR "nutritive"[All Fields]) OR "nutrition intervention*"[All Fields] OR "lifestyle*"[All Fields] OR "life style*"[All Fields] OR "feeding*"[All Fields] OR ("feeding behaviour"[All Fields] OR "feeding behavior"[MeSH Terms] OR ("feeding"[All Fields] AND "behavior"[All Fields]) OR "feeding behavior"[All Fields]) OR "sedentary behavior*"[All Fields] OR "sedentary behaviour*"[All Fields] OR ("sedentary behavior"[MeSH Terms] OR ("sedentary"[All Fields] AND "behavior"[All Fields]) OR "sedentary behavior"[All Fields] OR ("sedentary"[All Fields] AND "lifestyle"[All Fields]) OR "sedentary lifestyle"[All Fields]) OR "food*"[All Fields] OR ("food"[MeSH Terms] OR "food"[All Fields]) OR "food intake"[All Fields] OR ("eating"[MeSH Terms] OR "eating"[All Fields] OR ("food"[All Fields] AND "intake"[All Fields]) OR "food intake"[All Fields]) OR "food environment"[All Fields] OR "meal*"[All Fields] OR "dietary diversity"[All Fields] OR "fruit consumption"[All Fields] OR "fruit*"[All Fields] OR ("fruit"[MeSH Terms] OR "fruit"[All Fields] OR "fruits"[All Fields] OR "fruit s"[All Fields] OR "fruited"[All Fields] OR "fruiting"[All Fields]) OR "sugar*"[All Fields] OR ("sugar s"[All Fields] OR "sugared"[All Fields] OR "sugars"[MeSH Terms] OR "sugars"[All Fields] OR "sugar"[All Fields]) OR (("sugar s"[All Fields] OR "sugared"[All Fields] OR "sugars"[MeSH Terms] OR "sugars"[All Fields] OR "sugar"[All Fields]) AND ("intake"[All Fields] OR "intake s"[All Fields] OR "intakes"[All Fields])) OR "snack*"[All Fields] OR "sugar sweetened beverage*"[All Fields] OR "drink*"[All Fields] OR "fast food*"[All Fields] OR ("fast foods"[MeSH Terms] OR ("fast"[All Fields] AND "foods"[All Fields]) OR "fast foods"[All Fields] OR ("fast"[All Fields] AND "food"[All Fields]) OR "fast food"[All Fields]) OR "health behavior*"[All Fields] OR "health behaviour*"[All Fields] OR "unhealthy behavior*"[All Fields] OR "unhealthy behaviour*"[All Fields] OR "family-based"[All Fields] OR "community-based"[All Fields] OR "home-based"[All Fields] OR "school-based"[All Fields] OR "parent*"[All Fields] OR "teacher*"[All Fields] OR "active lesson*"[All Fields] OR "school lunch"[All Fields] OR ("lunchbox"[All Fields] OR "lunchboxes"[All Fields]) OR "lunch box"[All Fields] OR "school food"[All Fields] OR "tuckshop*"[All Fields] OR "vendor*"[All Fields] OR "food price*"[All Fields] OR "weight-related"[All Fields] OR "junk food"[All Fields] OR "screen time"[All Fields] OR "television viewing"[All Fields] OR (("televised"[All Fields] OR "televising"[All Fields] OR "television"[MeSH Terms] OR "television"[All Fields] OR "televisions"[All Fields] OR "television s"[All Fields]) AND ("viewed"[All Fields] OR "viewing"[All Fields] OR "viewings"[All Fields] OR "views"[All Fields])) OR "TV"[All Fields] OR "computer use"[All Fields] OR "portion size*"[All Fields] OR ("portion size"[MeSH Terms] OR ("portion"[All Fields] AND "size"[All Fields]) OR "portion size"[All Fields]) OR ("exergamers"[All Fields] OR "exergaming"[MeSH Terms] OR "exergaming"[All Fields] OR "exergame"[All Fields] OR "exergames"[All Fields]) OR "MVPA"[All Fields] OR "work-based"[All Fields] |
| Search term 3 | (("search"[All Fields] OR "searched"[All Fields] OR "searches"[All Fields] OR "searching"[All Fields] OR "searchs"[All Fields]) AND ("term birth"[MeSH Terms] OR ("term"[All Fields] AND "birth"[All Fields]) OR "term birth"[All Fields] OR "term"[All Fields]) AND "4"[All Fields] AND "Sub-Sahara Africa"[All Fields]) OR "West Africa"[All Fields] OR "East Africa"[All Fields] OR "Central Africa"[All Fields] OR "southern Africa"[All Fields] OR ("angola"[MeSH Terms] OR "angola"[All Fields] OR "angola s"[All Fields]) OR ("benin"[MeSH Terms] OR "benin"[All Fields] OR "benin s"[All Fields]) OR ("botswana"[MeSH Terms] OR "botswana"[All Fields] OR "botswana s"[All Fields]) OR "Burkina Faso"[All Fields] OR ("burundi"[MeSH Terms] OR "burundi"[All Fields]) OR ("cameroon"[MeSH Terms] OR "cameroon"[All Fields] OR "cameroons"[All Fields] OR "cameroon s"[All Fields]) OR "Cape Verde"[All Fields] OR "Central African Republic"[All Fields] OR ("chad"[MeSH Terms] OR "chad"[All Fields]) OR ("comoros"[MeSH Terms] OR "comoros"[All Fields] OR "comoro"[All Fields]) OR ("congo"[MeSH Terms] OR "congo"[All Fields]) OR "Democratic Republic of Congo"[All Fields] OR "DRC"[All Fields] OR ("djibouti"[MeSH Terms] OR "djibouti"[All Fields]) OR "Equatorial Guinea"[All Fields] OR ("eritrea"[MeSH Terms] OR "eritrea"[All Fields]) OR ("eswatini"[MeSH Terms] OR "eswatini"[All Fields]) OR ("ethiopia"[MeSH Terms] OR "ethiopia"[All Fields] OR "ethiopia s"[All Fields]) OR ("gabon"[MeSH Terms] OR "gabon"[All Fields]) OR ("gambia"[MeSH Terms] OR "gambia"[All Fields] OR "gambia s"[All Fields]) OR ("ghana"[MeSH Terms] OR "ghana"[All Fields] OR "ghana s"[All Fields]) OR ("guinea"[MeSH Terms] OR "guinea"[All Fields] OR "guinea s"[All Fields] OR "guineas"[All Fields]) OR "Guinea Bissau"[All Fields] OR "Ivory Coast"[All Fields] OR "Cote d'Ivoire"[All Fields] OR ("kenya"[MeSH Terms] OR "kenya"[All Fields] OR "kenya s"[All Fields]) OR ("lesotho"[MeSH Terms] OR "lesotho"[All Fields]) OR ("liberia"[MeSH Terms] OR "liberia"[All Fields] OR "liberia s"[All Fields]) OR ("madagascar"[MeSH Terms] OR "madagascar"[All Fields] OR "madagascar s"[All Fields]) OR ("malawi"[MeSH Terms] OR "malawi"[All Fields] OR "malawi s"[All Fields]) OR ("mali"[MeSH Terms] OR "mali"[All Fields]) OR ("mauritania"[MeSH Terms] OR "mauritania"[All Fields]) OR ("mauritius"[MeSH Terms] OR "mauritius"[All Fields]) OR ("mozambique"[MeSH Terms] OR "mozambique"[All Fields] OR "mozambique s"[All Fields]) OR ("namibia"[MeSH Terms] OR "namibia"[All Fields] OR "namibia s"[All Fields]) OR ("niger"[MeSH Terms] OR "niger"[All Fields]) OR ("nigeria"[MeSH Terms] OR "nigeria"[All Fields] OR "nigeria s"[All Fields]) OR "Republic of the congo"[All Fields] OR ("rwanda"[MeSH Terms] OR "rwanda"[All Fields] OR "rwanda s"[All Fields]) OR "Sao Tome and Principe"[All Fields] OR ("senegal"[MeSH Terms] OR "senegal"[All Fields] OR "senegal s"[All Fields]) OR ("seychelles"[MeSH Terms] OR "seychelles"[All Fields]) OR "Sierra Leone"[All Fields] OR ("somalia"[MeSH Terms] OR "somalia"[All Fields] OR "somalia s"[All Fields]) OR "South Sudan"[All Fields] OR ("sudan"[MeSH Terms] OR "sudan"[All Fields] OR "sudans"[All Fields] OR "sudan s"[All Fields]) OR ("tanzania"[MeSH Terms] OR "tanzania"[All Fields] OR "tanzania s"[All Fields]) OR ("togo"[MeSH Terms] OR "togo"[All Fields]) OR ("uganda"[MeSH Terms] OR "uganda"[All Fields] OR "uganda s"[All Fields]) OR ("zambia"[MeSH Terms] OR "zambia"[All Fields] OR "zambia s"[All Fields]) OR ("zimbabwe"[MeSH Terms] OR "zimbabwe"[All Fields] OR "zimbabwe s"[All Fields]) OR "Africa South of the Sahara"[MeSH Terms] |
